# “ICH-CARE”: *ICH*-integrated *C*are for *A*ccelerated *R*esponse to *H*emorrhage Using a Phased Approach

**DOI:** 10.64898/2026.07.18.26358392

**Authors:** Saif Salman, Stephen English, Lesia Mooney, David Miller, Lauren Ng, Christopher Kramer, Mutaz Ombada, Rabih Tawk, William David Freeman

**Affiliations:** Departments of Neurological Surgery, Neurology and Critical Care, Mayo Clinic, Jacksonville, Florida; Medical Director, Mayo Clinic Enterprise Telestroke Program; Co-Director, Comprehensive Stroke Center, Mayo Clinic, Jacksonville, Florida; Department of Nursing Mayo Clinic, Jacksonville, Florida

**Keywords:** Intracranial hemorrhage, subarachnoid hemorrhage, Patient care planning, thrombolytic therapy

## Abstract

**Introduction:** Intracerebral hemorrhage (ICH) carries higher morbidity and mortality than ischemic stroke. Recent studies have demonstrated improved patient outcomes by applying ultra-early bundled interventions including blood pressure management, coagulopathy reversal, and osmotic therapy. Effective strategies to deliver these ultra-early treatment options are currently being explored. On December 19th, 2022, the Mayo Clinic Comprehensive Stroke Center (CSC) launched the “ICH Phases” communication system to accelerate ICH patient care.

**Objective:** To evaluate adherence to the AHA/ASA guidelines in acute ICH care following the implementation of our novel-tiered paging system.

**Methods:** We retrospectively reviewed patients admitted with spontaneous ICH during 2024 and 2025. We excluded traumatic cases. We extracted clinical data such as time to imaging, documentation of ICH score, blood pressure control, reversal of anticoagulation, venous thrombo-embolism (VTE) prophylaxis and discharge disposition.

**Results:** Among 67 patients, 68.7% underwent CT imaging within 25 minutes. We documented the ICH score within 6 hours in 82.9% of patients. Nearly 94.7% of patients with SBP>140 mm Hg received antihypertensive therapy, yet only 18% reached target BP within 60 minutes. We completed the reversal of anticoagulation within 120 minutes in 75% of patients. VTE prophylaxis was initiated within 24 hours in 91% of patients.

**Discussion:** Our novel system demonstrated adherence to the AHA/ASA guidelines, and time sensitive benchmarks in neuroimaging, reversal of anticoagulation, and VTE prophylaxis. Early BP control remains a challenge, that highlights the discrepancy between guidelines and real-ground implementation.

**Conclusion:** A novel tiered paging system is effective for enhancing early ICH care. Such a holistic system remains critical for sustained improvement in quality of care.

## Introduction

Intracranial hemorrhage (ICH) is a disproportionately deadly and disabling subtype of stroke. While the ischemic subtype is more common, ICH surpasses it in terms of mortality and morbidity with an incidence of 73/100,000 per year, and thirty-day mortality as high as 40% that can reach up to 60% in one year^1^. ICH is a medical emergency that requires rapid intervention. Acute interventions to lower blood pressure and reverse coagulopathy are critically important, as hypertension and anticoagulant use potentiate the risk of hematoma expansion and rebleeding.

Hence, minimizing hematoma expansion is pivotal in controlling *ICH*^2^ and ensuring safe and timely surgical interventions^3^. The INTERACT3 trial shows a promising role in utilizing a bundled approach to ICH management. This includes the hyperacute bundle of care for reversal of anticoagulation, intensive lowering of blood pressure, neurosurgical assessment, and facilitated access to NSICU^2,4,5^.

The updated American Heart Association (AHA)/American Stroke Association (ASA) guidelines for ICH^6^ emphasize early diagnosis as a crucial step to promote timely intervention beginning from the pre-hospital phase. Public education and training of the first responders to recognize presenting signs and symptoms, notifying the hospital before arrival, and facilitating transportation, can ensure a subsequent expedited in-hospital response, and a shorter time for imaging and intervention. This is fortified by a holistic system that provides rapid mobilization of resources and care teams^6–17,^.

The AHA/ASA ICH guidelines further emphasize expedited imaging to differentiate ICH from ischemic stroke and determine the hemorrhage volume. Neuroimaging is mandatory to confirm the diagnosis. This is followed by immediate notification of the ICH team including neurosurgery, neurocritical care, emergency medicine, stroke, and pharmacy teams. Prioritizing blood pressure control using IV clevidipine, nicardipine, labetalol, and hydralazine, alongside urgent correction of coagulopathy should be targeted within 60 minutes of arrival^2,6–8^. If systolic blood pressure (SBP) ranges between 150 to 220 mm Hg, then it is safe to lower it to 140 mm Hg. However, if SBP exceeds 220 mm Hg then aggressive BP lowering with continuous infusion is recommended.

Expedited laboratory testing is crucial to uncover underlying coagulopathies, electrolyte disturbances, and organ dysfunction. Notably, coagulation status affects the choice of intervention in due course. However, when considering the reversal of anticoagulation, medical history is superior to laboratory testing. The 2024 AHA/ASA quality measures emphasize that anticoagulation reversal should occur as rapidly as possible, ideally initiated immediately upon identification of anticoagulant-related ICH, to prevent the hematoma expansion as a delayed sequalae Prothrombin complex concentrates (PCC) are preferred over fresh frozen plasma (FFP) for the reversal of warfarin-induced anticoagulation. Idarucizumab or Andexanet alfa are recommended in cases involving direct oral anticoagulants^6,11–17^.

The decision for an expedited surgical intervention begins in the ED. Surgical intervention is considered in deteriorating patients with lobar hemorrhage, cerebellar hemorrhage that exceeds 3 cm, or with brainstem compression^6,18,19^. Minimally invasive surgery is considered in some cases but that is beyond the scope of this manuscript.

External ventricular drains (EVD) and intracranial pressure (ICP) monitoring are considered in cases with hydrocephalus or brain herniation^6, 20, 21^. Electroencephalogram (EEG) is recommended in non-convulsive seizures. Prophylactic anticonvulsants are not recommended in the absence of seizures^6^.

Pneumatic compression should be started on admission as a prophylaxis for deep vein thrombosis (DVT). Once bleeding is stabilized and the risk of DVT is high, then LMWH can be considered later on.

On December 19^th^, 2022, the Mayo Clinic Comprehensive Stroke Center (CSC) launched the “ICH Phases” communication system to accelerate ICH patient care. This progressive system encompasses a pre-hospital phase, an ED phase (hospital arrival to ICH identification on Non-contrast CT (NCCT) scan), and a hospital phase that includes rapid multidisciplinary evaluation by vascular neurology, neurocritical care, interventional neuroradiology, and neurosurgery.

In this study, we describe our phased tiered paging system-of-care developed to facilitate ultra-early interventions for patients with ICH.

## Methods

We conducted a retrospective review for patients admitted to our CSC from 2024 to 2025 with intracerebral hemorrhage (ICH). Patients were identified based on ICD-10 codes and a review of medical reports. We excluded traumatic cases, and transfers from other hospitals due to incomplete documentation. We extracted clinical data from our electronic medical records such as demographics, arrival mode, blood pressure parameters, use of anticoagulants, imaging and treatment timeframes, and discharge medications. Further subclassification included door-to-CT time, time-to-ICH score documentation, time to BP management initiation, time to anticoagulation reversal, initiation of prophylaxis for venous thromboembolism, and discharge on antihypertensive medications.

Our primary objective was to assess adherence to benchmarks based on the AHA/ASA acute ICH management measures^6^. Descriptive analysis was used to deduce adherence to performance measures.

## Results

After exclusion, a total of 67 patients with spontaneous ICH were included. Their median age was 74 years.

### Timing of neuroimaging and diagnosis

Of the 67 patients, 46 patients (68.7%) underwent CT scanning within 25 minutes of arrival, meeting the benchmark for door-to-imaging time. We documented the ICH score in 63 patients (82.9%) within 6 hours of arrival.

ICH score was documented in 82.9% of our patients within 6 hours of arrival. Our institutional target was to achieve 90%.

### Blood pressure measurement

Of the 67 patients, 56 patients (83.6%) had SBP >140 mm. Among these, 53 patients (94.7%) received antihypertensive medications. Further, among the latter, 27 patients (48.2%), 38 patients (67.9%), and 43 patients (76.8%) received antihypertensive medications on 30, 45, and 60 minutes, respectively. BP control to target thresholds was achieved in 18%, 57%, and 71% by 60, 90, and 120 minutes, respectively.

### Anticoagulation reversal

Of the 67 patients, 16 (21.1%) received anticoagulation at presentation. Drug-specific anticoagulation reversal within 120 minutes was achieved in 12 patients (75.0%). Reversal occurred within 60 minutes in 3 patients (18.8%) and within 90 minutes in 9 patients (56.3%). Failure to undergo reversal in the remaining 4 patients (25.0%) was attributed to subtherapeutic INR, delayed timing since the last anticoagulant dose, or subacute presentation.

### VTE prophylaxis

Of the 67 patients, compression devices were used in 61 patients (91%) within 24 hours.

### Discharge recommendations

Of the 67 patients, 18 (26.9%) died during hospitalization. Among the 49 patients discharged alive, 41 (83.7%) were discharged on antihypertensive medications, while the remaining 8 patients were normotensive and did not require antihypertensive medications.

## Discussion

We implemented a novel ICH Phases paging system in December 2022 to accelerate response in our ICH treatment teams (Figure 1). The novelty in our system resides in the holistic engagement of multidisciplinary teams, enhancing communication, promoting intervention in an ultra-timely manner, and the progressive establishment of target goals and benchmarks that align with the proposed performance measures from the AHA/ASA.

**Figure 1:**
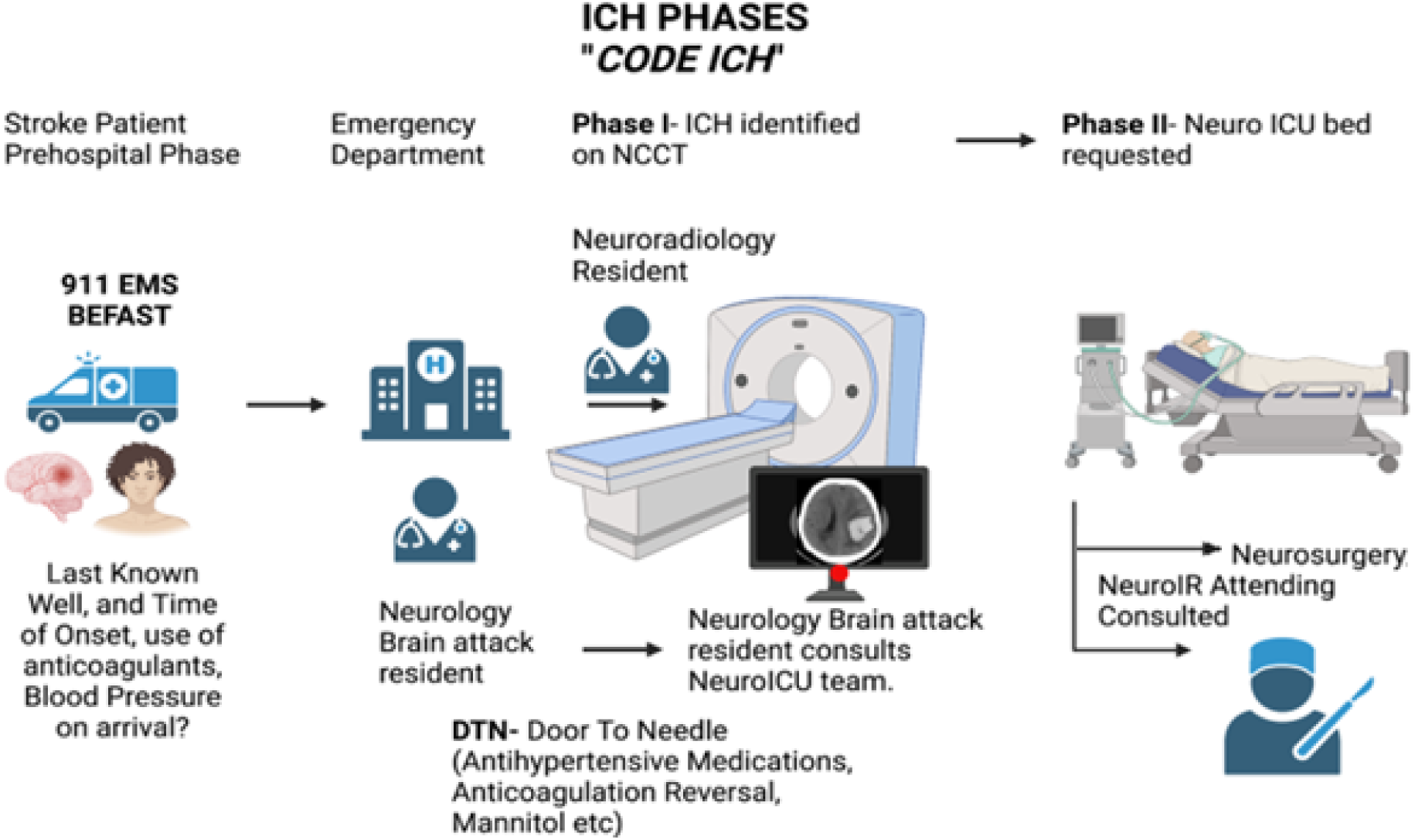
Visual Flow Diagram for “ICH CARE” Phases of communication system launched by the Mayo Clinic Comprehensive Stroke Center (CSC).

The hospital workflow begins with ED admission and identification of ICH on non-contrast CT (NCCT) scans by the neurology resident. If the patient is receiving anticoagulation therapy, has an ICH score greater than 1, or presents with systolic blood pressure greater than 160 mmHg, phase 2 of the care pathway is activated to mobilize the NSICU and neurosurgery teams and expedite early targeted medical and surgical interventions. These interventions include aggressive blood pressure control with intravenous clevidipine, reversal of anticoagulation, and evaluation for EVD placement, minimally invasive hematoma evacuation, or decompressive hemicraniectomy.

Timely neuroimaging is crucial for the acute management in patients with ICH. In our study, 69% of patients underwent imaging within 25 minutes of arrival. Although promosing, this remains below our institutional targets and highlights the need for a more streamlined imaging workflow. Importantly, several barriers unique to hemorrhagic stroke may contribute to delays compared with ischemic stroke workflows. Patients with primary IVH or SAH may present without focal neurological deficits, resulting in delayed recognition and triage. In addition, patients with hemorrhagic stroke require early airway stabilization, hemodynamic management, or treatment of elevated intracranial pressure prior to CT acquisition, which may prolong imaging timelines. Future integration of AI-driven imaging and triage tools may help overcome these challenges by accelerating hemorrhage detection, automating imaging interpretation, and facilitating earlier neurosurgical mobilization and intervention.^22,23^ (Figure 2)

**Figure 2:**
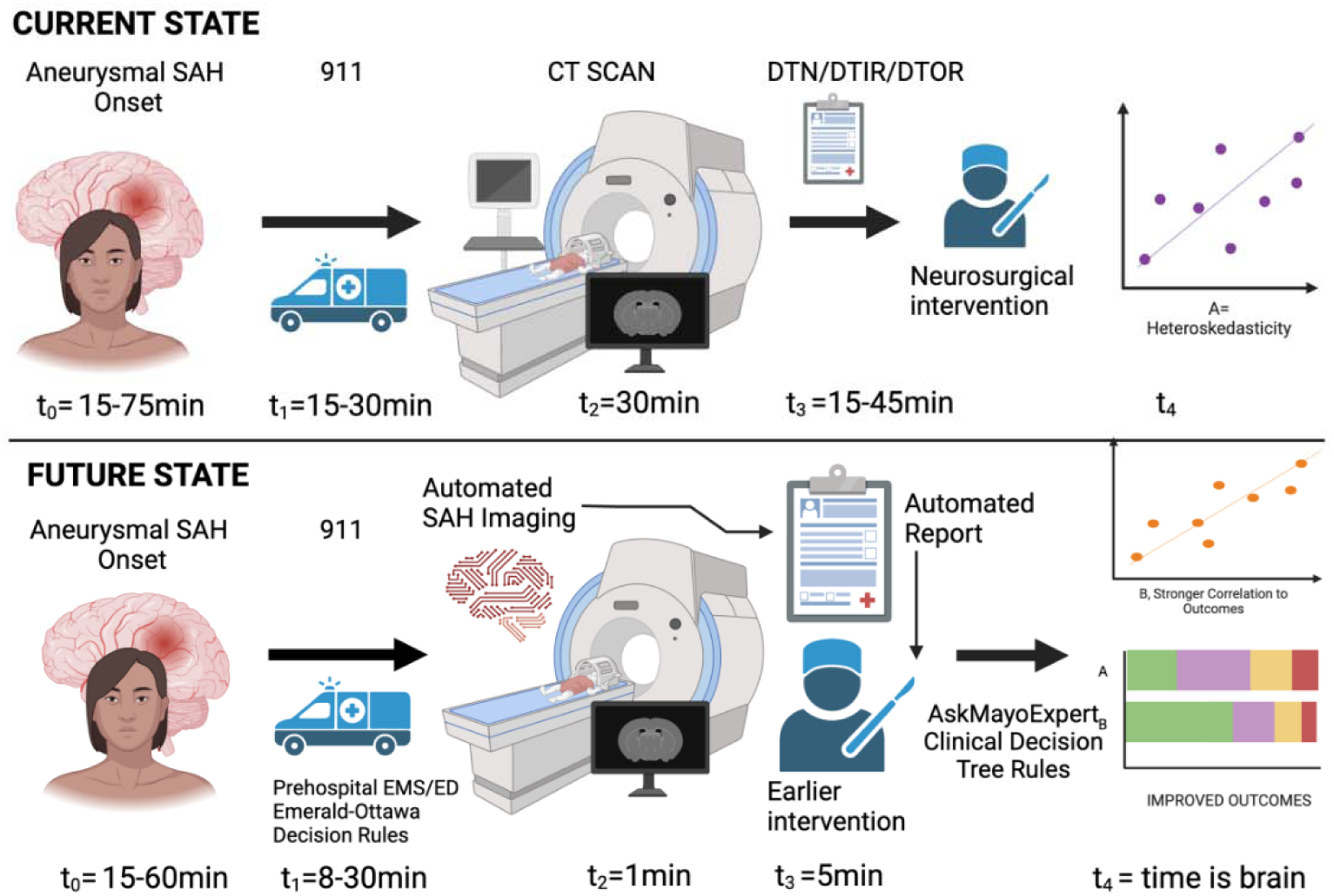
Representation of a future framework wherein AI-driven tools optimize imaging and interpretation in a timely manner.

Implementation of validated prognostic tools such as the ICH score in the ED setting may facilitate shared decision-making with families, early palliative care consultation, and disposition planning, including acute inpatient rehabilitation or transfer to rehabilitation facilities. As ICH score documentation represents an established AHA/ASA quality metric, continued efforts to improve compliance through standardized checklists and workflow integration may further enhance consistency during the initial evaluation phase^24–26^.

Blood pressure control within the first hour is associated with reduced hematoma expansion. In our cohort, antihypertensive therapy was administered in 94.7% of patients with SBP >140 mmHg. Among these patients, antihypertensive treatment was initiated within 30, 45, and 60 minutes in 48.2%, 67.9%, and 76.8% of cases, respectively. However, target blood pressure thresholds were achieved in only 18% of patients by 60 minutes. In correlation with the INTERACT-3 trial, it is difficult to achieve early BP control in real hospital settings^27^.

In our cohort, anticoagulation reversal within the AHA/ASA-recommended 120-minute benchmark was achieved in 75% of anticoagulated patients. However, only 18.8% of patients underwent reversal within 60 minutes, highlighting persistent delays in hyperacute anticoagulation management. We believe these delays primarily reflect challenges in early decision-making and logistical coordination, particularly in cases with unclear medication history, delayed medication reconciliation, or uncertain anticoagulation status at presentation. Standardized reversal protocols and enhanced prehospital medication reconciliation may further improve adherence to ultra-early reversal targets^28^.

We successfully initiated VTE prophylaxis in 91% of patients within 24 hours exceeding our designated institutional target. This emphasizes the role of implicating evidence-based medicine into care systems^29^. Discharge settings from the hospital include managing risk factors such as hypertension, early mobilization and rehabilitation, and scheduling future appointments. In our study, the rate of prescriptions for antihypertensive medications at discharge approached 84%. This reflects the continuity in managing risk factors. However, we still believe in chances for improvement.

A major limitation in our study is the descriptive analysis and retrospective nature in this single-center design. A multicenter study is needed in the future.

## Conclusion

ICH systems of care should be approached holistically. Adoption of a phased multidisciplinary paging system may be beneficial in facilitating ultra-early interventions and improving adherence to time-sensitive quality measures in patients with ICH.

## Data Availability

All data produced in the present study are available upon reasonable request to the authors

